# Are culturally vibrant communities healthier? Relationships between performing arts activity and health outcomes in the 500 largest US cities

**DOI:** 10.1101/2021.07.26.21261123

**Authors:** J. Matt McCrary, Michael Großbach, Eckart Altenmüller

## Abstract

**Aims:** Recent reviews have demonstrated broad links between performing arts participation (*e.g. music-making; dancing; acting*) and receptive engagement (*e.g. listening to music; attending a dance/theatre performance)* and improved health, including reduced disease and mortality risk. However, no investigations to date have interrogated the links between community-level performing arts activity (*i.e. participation + receptive engagement*) and health outcomes – i.e. do the performing arts help create healthy communities? This study aims to address this question by examining links between performing arts activity and health outcomes across 500 cities in the United States.

**Methods:** Secondary analysis of demographic, health outcome, performing arts activity (estimated by annual performing arts revenue), and preventive/unhealthy behaviour data for 500 large cities in the United States – data extracted from the US Centers for Disease Control 500 Cities Project, Dun & Bradstreet Hoovers Database, and US Census. Links between performing arts activity and 12 health/disease outcomes were evaluated using a series of hierarchical beta regression models which progressively controlled for demographic variables and preventive/unhealthy behaviour prevalence.

**Results:** The 500 analysed US cities comprise 33.4% of the total US population and 84,010 performing arts businesses (total annual revenue $27.84 billion). No significant associations were found between performing arts activity and nine of twelve health outcomes in fully adjusted models (p≥.17). Statistically significant relationships (p<.01) between increased performing arts activity and increased prevalence of chronic kidney disease, coronary heart disease, and stroke were determined to be clinically equivocal.

**Conclusions:** This study contributes to a growing body of conflicting epidemiologic evidence regarding the impact of the performing arts on health/disease and mortality outcomes, evaluated using a range of disparate methodologies. A consensus, psychometrically rigorous approach is required to address this prevailing uncertainty in future epidemiologic studies examining effects of performing arts activities both within and across countries and communities.

## Introduction

Recent systematic and scoping reviews have demonstrated broad links between performing arts participation (*e.g. music-making; dancing; acting*) and receptive engagement (*e.g. listening to music; attending a dance or theatre performance)* and improved health and wellbeing (1, 2). Included amongst these broad health benefits are suggestive links between the performing arts and a reduced risk of early mortality and non-communicable diseases including cancer, dementia and cardiovascular disease (3-7). However, no investigations to date have interrogated the links between community-level performing arts activity (*i.e. participation + receptive engagement*) and health outcomes – in other words, *do the performing arts help create healthy communities?*

In addition to being directly health-promoting activities, the performing arts are also substantial contributors to culture in communities in the United States, with culture also independently noted to substantially impact health (8). The performing arts (*i.e. music, dance, theatre*) are the most popular artistic modalities in the United States - 43% of adults attend performing arts events at least once annually; 74% engage with the performing arts using electronic media (*e.g. television; online*); and 40% participate in the performing arts (*i.e. play an instrument, sing, dance, act*) at least once every year (9). This study aims to provide insights into the impact of the performing arts on community health by examining links between performing arts activity and health outcomes across 500 cities in the United States.

## Methods

Study aims were addressed through secondary analyses of data obtained from the US Centers for Disease Control and Prevention 500 Cities Project (10), Dun & Bradstreet Hoovers Database (11), and the US Census Bureau (12).

### US Centers for Disease Control and Prevention 500 Cities Project (10) – Health Outcomes; Unhealthy & Preventive Behaviours

The 500 Cities Project uses small area estimation methods (13) to approximate the prevalence of 13 health outcomes, 9 preventive behaviours, and 5 unhealthy behaviours in the 497 largest US cities (as per 2010 US census). Additionally, to ensure representation from each US state, data from the largest cities in Vermont, West Virginia and Wyoming are included. The 2019 release of 500 Cities Project data was used in the present study.

Included health outcomes are: *arthritis; asthma; cancer; chronic kidney disease; chronic obstructive pulmonary disease (COPD); coronary heart disease; diabetes; high blood pressure; high cholesterol; poor mental health; poor physical health; tooth loss (all teeth); and stroke*. Included unhealthy behaviours are: *binge drinking; smoking, physical activity; obesity; and insufficient sleep*. Included preventive behaviours are (restricted to appropriate age groups as relevant): *annual dental visit; annual medical check-up; breast cancer screening (mammography); cervical cancer screening (Papanicolanou smear); cholesterol screening; colorectal cancer screening (colonoscopy, sigmoidoscopy, faecal occult blood test); core set of clinical preventive services for older adults; health insurance coverage; and high blood pressure medication adherence*. All data are reported as an age-adjusted % prevalence of the respective health outcome or behaviour for each of the 500 included cities. Full details of the 500 Cities Project can be found in (10).

### Dun & Bradstreet Hoovers (11) – Performing Arts Activity

Performing arts activity was estimated for each of the 500 cities included in the 500 Cities Project by extracting annual revenue (2019) data for performing arts-related for- and non-profit entities from the Dun & Bradstreet Hoovers database. Hoovers is the world’s largest commercial financial database including over 170 million businesses, and has been previously used to estimate cultural activity in US cities and analyse relationships between cultural activity and health outcomes (14). Performing arts-related entities were identified by eight-digit Standard Industrial Classification (SIC) codes (15), with 78 codes related to performing arts participation and/or receptive engagement selected for inclusion (Supplementary Table 1). All included SIC codes are related to participation and/or receptive engagement with live performing arts. SIC codes related to recorded performing arts were explicitly excluded given more tenuous links between revenue and performing arts participation/receptive engagement.

### US Census Bureau (12) – Demographic covariates

Population, median per capita income and racial (% White, African-American, Hispanic) data were extracted from the most recent US Census (2010) for each of the 500 cities included in the 500 Cities Project. Demographic data were extracted to serve as covariates given the demonstrated impact of economic and racial disparities on health outcomes (16, 17).

### Statistical analyses

Relationships between performing arts activity and each health outcome were analysed using a series of hierarchical beta regression models. Beta regression has been shown to be ideal for effectively modelling proportion outcomes (*i.e. the 13 included health outcomes*) which are limited to the interval [0,1](18). Hierarchical modelling enabled consideration of established social and economic relationships between cities contained within the same metropolitan area; for example, hierarchical models could appropriately treat Santa Monica, Los Angeles (city), and Long Beach as related cities with the Los Angeles metropolitan area, rather than completely independent entities (19). Hierarchical models included two levels: Level 1 = metropolitan areas including at least one of the analysed 500 cities; Level 2 = the individual 500 cities, coded and stratified by metropolitan area. Metropolitan areas were defined using Combined Statistical Area designations assigned by the US Office of Management and Budget in reflection of social and economic links between cities (20, 21).

Four hierarchical beta regression models were created using the *PROC GLIMMIX* procedure in SAS v9.4 (SAS Institute Inc., Cary, NC, USA) to analyse the relations between performing arts activity and each of the 13 health outcomes across the 500 included cities:

Model 1 – completely unadjusted

Model 2 – adjusted for demographic covariates (*median annual per capita income; population size; % White; % African-American; % Hispanic*)

Model 3 – adjusted for everything in Model 2 + prevalence of five unhealthy behaviours

Model 4 – adjusted for everything in Model 3 + prevalence of nine preventive behaviours

All models exceed recommended ratios of observation to predictor variables; additionally, a priori designation of covariates has been shown to minimize potential risks of overfitting (22). Variance Inflation Factors for predictor variables (*i.e. performing arts activity*) were checked to ensure that potentially problematic levels of multicollinearity (Variance Inflation Factor > 10) were not present (23). Using the logit link, odds ratios were derived from parameter estimates of each model (18); odds ratios are multipliers describing the impact of a $1 billion increase in performing arts revenue on the % prevalence of each health outcome. Analyses of cancer prevalence were excluded due to particularly tight clustering of cancer prevalence data which precluded accurate modelling. Missing cervical cancer screening data from 47 cities were multiply imputed (10 imputations) using the *PROC MI* procedure (SAS v9.4); Model 4 parameter estimates were averaged across the 10 multiply imputed datasets using the *PROC MIANALYZE* procedure (SAS v9.4). Data for all other investigated variables were complete.

## Results

The 500 analysed US cities comprise 33.4% of the total U.S. population (103,020,808 people) and 84,010 performing arts businesses with a total annual revenue of $27.84 billion. Of this total performing arts revenue, $10.42 billion went to performing artists, $11.84 billion went to performing arts production/support services, $3.30 billion went to performing arts venues, and $2.27 billion went to performing arts education. Median, minimum and maximum values for demographic and health and behavioural outcomes across the 500 analysed cities are detailed in Table 1.

**Table 1.** Descriptive demographic, health outcome and behavioural data for the 500 analysed cities.

|  | Median | Minimum | Maximum |
| --- | --- | --- | --- |
| Annual performing arts revenue (\$ - millions) | 7.56 | 0.29 | 4186.22 |
| Population | 106,106 | 42,417 | 8,175,133 |
| Median annual per capita income (\$) | 29,031 | 14,509 | 90,042 |
| ***Racial Demographics (%)*** |  |  |  |
| White | 72.2 | 12.3 | 96.8 |
| African-American | 9.7 | 0.0 | 84.9 |
| Hispanic | 16.4 | 1.0 | 96.8 |
| ***Health Outcomes (% prevalence)*** |  |  |  |
| Arthritis | 22.0 | 13.3 | 33.9 |
| Asthma | 9.4 | 6.7 | 14.2 |
| Chronic kidney disease | 3.1 | 2.1 | 4.8 |
| Chronic obstructive pulmonary disease | 6.1 | 3.1 | 11.3 |
| Coronary heart disease | 5.7 | 3.5 | 8.8 |
| Diabetes | 10.3 | 5.6 | 20.3 |
| High blood pressure | 30.0 | 20.7 | 47.3 |
| High cholesterol | 29.5 | 24.1 | 34.1 |
| Poor mental health | 13.6 | 8.3 | 19.6 |
| Poor physical health | 12.7 | 7.0 | 20.5 |
| Teeth lost (all) | 14.5 | 5.1 | 31.8 |
| Stroke | 3.1 | 1.8 | 6.1 |
| ***Unhealthy Behaviours (% prevalence)*** |  |  |  |
| Binge drinking | 17.7 | 6.2 | 25.4 |
| Current smoker | 17.2 | 7.9 | 29.6 |
| Insufficient sleep | 35.4 | 24.5 | 50.1 |
| Physical inactivity | 26.2 | 12.9 | 45.4 |
| Obesity | 30.5 | 15.3 | 49.1 |
| ***Preventive Behaviours (% prevalence)*** |  |  |  |
| Annual dental | 63.4 | 41.8 | 81.5 |
| Annual medical | 67.9 | 54.2 | 81.3 |
| Breast cancer screening | 74.9 | 60.0 | 83.5 |
| Cervical cancer screening | 78.6 | 67.7 | 85.9 |
| Cholesterol screening | 79.8 | 70.2 | 85.2 |
| Colon cancer screening | 64.6 | 43.2 | 77.7 |
| Health insurance | 15.1 | 4.8 | 43.9 |
| High blood pressure medication adherence | 57.1 | 46.6 | 69.7 |
| Preventive services – older men | 34.0 | 19.4 | 53.3 |
| Preventive services – older women | 32.1 | 17.5 | 46.6 |

Fully adjusted regression models (Model 4) demonstrate statistically significant associations between increased performing arts activity and increased prevalence of chronic kidney disease, coronary heart disease, and stroke (p<.01; Table 2). Odds ratios indicate that a $1 billion increase in performing arts revenue is linked to increases in chronic kidney disease, coronary heart disease, and stroke by factors of 1.06, 1.13, and 1.11, respectively (Figure 1). For a hypothetical city of median population, performing arts revenue and incidence of health outcomes, Model 4 demonstrates that a 100% increase in performing arts revenue would be associated with an additional 2, 6, and 3 cases of chronic kidney disease, coronary heart disease, and stroke, respectively. No other significant links between performing arts activity and health outcomes were found in fully adjusted models (Model 4; p≥.17).

**Table 2.** Hierarchical beta regression results describing the relationships between performing arts activity and all health outcomes * - p<.05: ** - p<.01; p<.001. Model 1: Unadjusted. Model 2: Adjusted for population size, median annual per capita income, and racial demographics (% White; % African-American; % Hispanic). Model 3: Adjusted for all in Model 2 + prevalence of unhealthy behaviours. Model 4: Adjusted for all in Model 3 + prevalence of preventive behaviours.

|  | Model 1 |  |  | Model 2 |  |  | Model 3 |  |  | Model 4 |  |  |
| --- | --- | --- | --- | --- | --- | --- | --- | --- | --- | --- | --- | --- |
| Outcome | Parameter estimate | SE | p | Parameter estimate | SE | p | Parameter estimate | SE | p | Parameter estimate | SE | p |
| Arthritis | -0.045 | 0.039 | 0.25 | 0.062 | 0.059 | 0.29 | 0.011 | 0.039 | 0.78 | 0.009 | 0.037 | 0.81 |
| Asthma | 0.008 | 0.026 | 0.77 | 0.065 | 0.053 | 0.22 | 0.034 | 0.043 | 0.43 | -0.018 | 0.037 | 0.53 |
| Chronic kidney disease | 0.047 | 0.027 | 0.09 | 0.103** | 0.035 | <0.01 | 0.061* | 0.027 | 0.02 | 0.061** | 0.023 | <0.01 |
| Chronic obstructive pulmonary disease | -0.00001 | 0.050 | 0.99 | 0.225** | 0.081 | <0.01 | 0.063 | 0.046 | 0.17 | 0.059 | 0.042 | 0.17 |
| Coronary heart disease | 0.025 | 0.034 | 0.47 | 0.207*** | 0.052 | <0.001 | 0.115*** | 0.024 | <0.001 | 0.120*** | 0.023 | <0.001 |
| Diabetes | 0.077 | 0.044 | 0.08 | 0.057 | 0.063 | 0.37 | 0.001 | 0.036 | 0.98 | 0.036 | 0.033 | 0.28 |
| High blood pressure | 0.021 | 0.042 | 0.62 | -0.029 | 0.061 | 0.63 | -0.069 | 0.039 | 0.08 | -0.012 | 0.035 | 0.73 |
| High cholesterol | 0.009 | 0.018 | 0.60 | 0.006 | 0.041 | 0.88 | -0.017 | 0.035 | 0.62 | 0.022 | 0.032 | 0.51 |
| Poor mental health | -0.001 | 0.036 | 0.97 | 0.169* | 0.055 | 0.02 | 0.076* | 0.035 | 0.03 | 0.041 | 0.031 | 0.19 |
| Poor physical health | 0.022 | 0.042 | 0.60 | 0.188** | 0.059 | <0.01 | 0.081* | 0.034 | 0.02 | 0.043 | 0.029 | 0.14 |
| Teeth lost (all) | 0.071 | 0.067 | 0.29 | 0.376*** | 0.113 | <0.001 | 0.177* | 0.078 | 0.02 | 0.041 | 0.063 | 0.52 |
| Stroke | 0.064 | 0.037 | 0.09 | 0.204*** | 0.045 | <0.001 | 0.122*** | 0.030 | <0.001 | 0.102*** | 0.025 | <0.001 |

**Figure 1.**
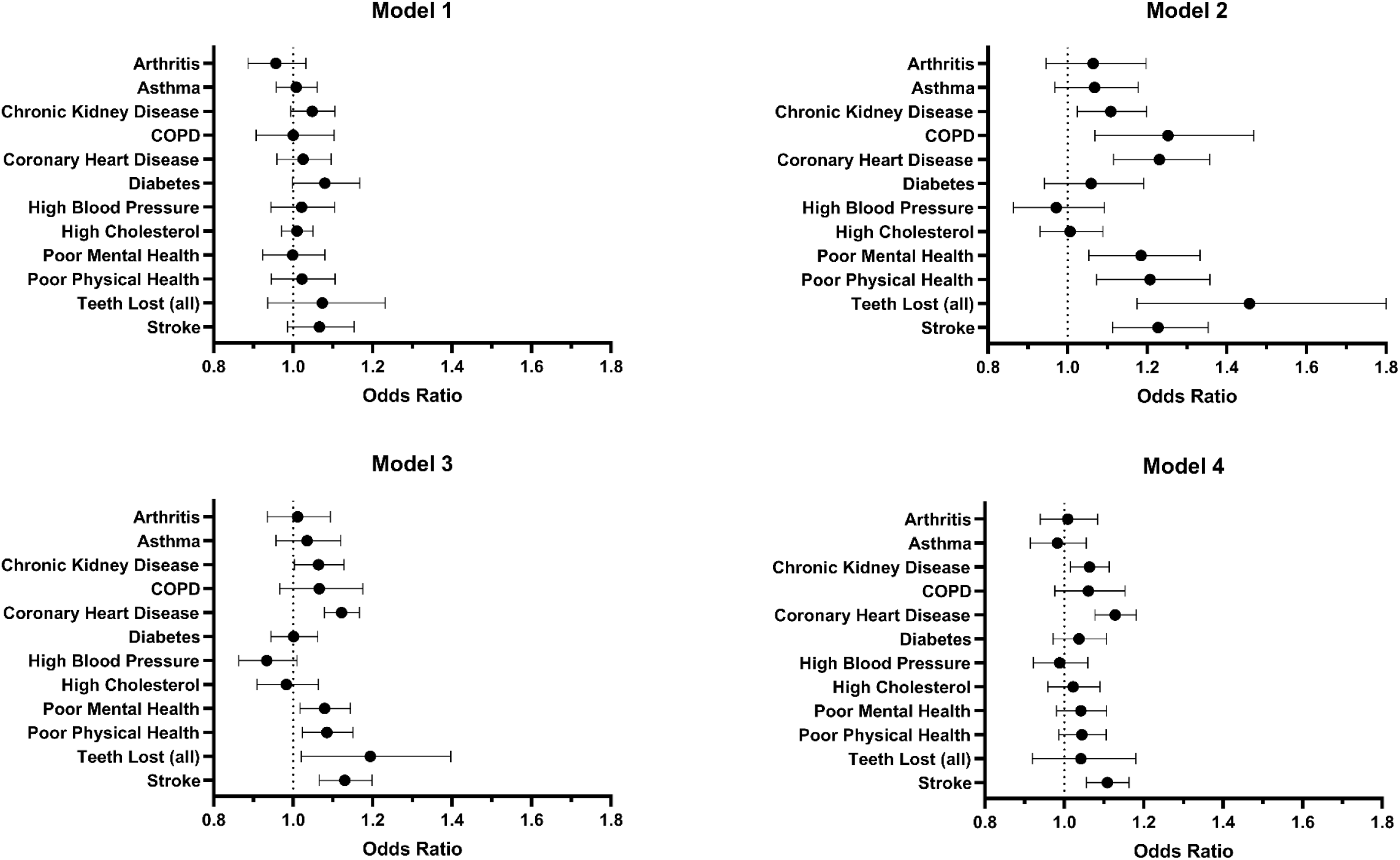
Odds ratios (± 95% confidence interval) generated from hierarchical beta regression results using the logit link (18). Odds ratios are multipliers describing the impact of a $1 billion increase in performing arts revenue on the % prevalence of each Health Outcome.

## Discussion

This secondary analysis of performing arts activity and health outcome data across 500 US cities representing 1/3 of the total US population revealed minimal links between community-level performing arts activity and health outcomes. Fully adjusted models only revealed statistically significant relationships between performing arts activity and an increased incidence of three of twelve investigated health outcomes: chronic kidney disease, coronary heart disease, and stroke incidence. However, the public health implications of these statistically significant links are equivocal – models indicate that considerable 100% increases in performing arts revenue would be linked to only single digit increases in disease incidence in most analysed cities. Such small but statistically significant associations in observational epidemiologic research have been shown to be most likely the result of uncontrolled bias and thus unlikely to be credible (24-26). Further, performing arts participation has been shown to positively impact chronic kidney disease, coronary heart disease and stroke risk factors (*e.g. body composition; inflammation*) in prior studies (1); no links between performing arts activity and increased incidence or risk factors for these diseases have been reported (1, 2). Accordingly, the results of this analysis are judged to illustrate an absence of *clinically* significant associations between performing arts activity and health outcomes – the remainder of the discussion has been written on this basis.

Given the broad benefits of performing arts participation and receptive engagement related to a range of health outcomes (1, 2), the absence of links between community-level performing arts activity and health outcomes in the present study was unexpected. However, null results of the present study are far from the exception in epidemiologic studies investigating performing arts impact. Only one of three studies found a positive effect of performing arts activity on all-cause mortality (5, 27, 28), while two of three studies have demonstrated protective effects of performing arts activity on dementia incidence (3, 7, 29). Methods of evaluating performing arts participation and receptive engagement vary across this and prior epidemiologic studies, with a notable dearth of validated and/or psychometrically tested approaches. Taken together, variation in the content and psychometric rigour of evaluation methods seems likely to be at least partially responsible for the present prevalence of mixed results.

In the present study, use of annual performing arts revenue to estimate performing arts activity is a notably indirect assessment approach and a key limitation. The authors hypothesized that the health impact of the performing arts would be robust enough to tolerate such indirect estimation methods, particularly given the availability of a wide array of behavioural and demographic covariates which would theoretically permit the detection of smaller effects. However, results indicate that the effects of performing arts activity may be smaller and/or more specific than predicted. Additionally, associations between performing arts revenue and participation/receptive engagement may be weaker than prior research (14) suggests. And alternately, performing arts activity may simply have no effect on broad health/disease outcomes. Further epidemiologic research using a consensus, psychometrically tested approach to evaluating performing arts activity is required to more confidently and precisely quantify performing arts effects.

Notably, studies which more directly assessed performing arts activity – e.g. questionnaires regarding the type and/or frequency of performing arts activities – have also returned conflicting null and positive results (3, 5, 7, 27-29). This suggests that direct assessment methods are not inherently the solution. However, one prior study interrogating links between dancing and cardiovascular disease mortality provides guidance regarding an approach that could be expanded to evaluate performing arts more broadly (6). This study used a validated interviewer-administered questionnaire to quantify the frequency, duration *and* physical intensity of dancing, finding that moderate, but not light, intensity dancing was linked to reduced cardiovascular disease mortality.

While physical intensity is not relevant to many forms of performing arts participation and receptive engagement, direct assessment of the physiologic response to performing arts activity appears likely to facilitate better evaluations of performing arts’ impact on health outcomes. Physiologic responses to performing arts have been shown to be highly variable, even within the same type of performing arts participation or receptive engagement – in a particularly clear example, performing or listening to the same piece of music can elicit a significant physiologic response in one individual but no response in another, both on average and during emotional ‘peaks’ in the music (1, 30-35). Evidence from other domains, in particular physical activity, has demonstrated clear links between the short-term physiologic responses to activities and their long-term impact on disease and mortality (36, 37). Similar links between short-term physiologic responses to performing arts activities and long-term health outcomes seem likely, underscoring the prospective importance of evaluating these physiologic responses in future epidemiologic research.

## Conclusions

This study contributes to a growing body of conflicting epidemiologic evidence regarding the health impacts of performing arts activity, revealing no relationships between community-level performing arts activity and health/disease outcome across 500 cities in the US. However, considering the context, results should be interpreted as an impetus to refine and consolidate presently disparate evaluation methods, rather than as conclusive insights regarding the impact of performing arts activity on community-level health. A consensus, psychometrically rigorous method of evaluating performing arts participation and receptive engagement is required to address this prevailing uncertainty in future epidemiologic studies.

## Data Availability

The full dataset used in analyses described in this manuscript will be uploaded alongside the final, peer-reviewed manuscript.

