## Supplementarz Table 1 for "Are culturally vibrant communities healthier? Relationships between performing arts activity and health outcomes in the 500 largest US cities"

| **Category** | **8-digit SIC Code** | **Code Description** |
| --- | --- | --- |
| Performing artists | 79220300 | Theatrical companies |
|  | 79220301 | Amateur theatrical company |
|  | 79220302 | Burlesque company |
|  | 79220303 | Opera company |
|  | 79220304 | Plays, road and stock companies |
|  | 79220305 | Repertory, road or stock companies: Theatrical |
|  | 79220306 | Summer theater |
|  | 79290000 | Entertainers and entertainment groups |
|  | 79290100 | Musical entertainers |
|  | 79290101 | Chamber music groups or artists |
|  | 79290102 | Classical music groups or artists |
|  | 79290103 | Country music groups or artists |
|  | 79290104 | Dance band |
|  | 79290105 | Drum and bugle corps (drill teams) |
|  | 79290106 | Gospel singers |
|  | 79290107 | Jazz music group or artists |
|  | 79290108 | Musician |
|  | 79290109 | Orchestras or bands, NEC |
|  | 79290110 | Popular music groups or artists |
|  | 79290111 | Symphony orchestra |
|  | 79299901 | Actor |
|  | 79299902 | Actress |
|  | 79299903 | Disc jockey service |
|  | 79299904 | Entertainers |
|  | 79299905 | Entertainment group |
|  | 79299906 | Entertainment service |
|  | 79299907 | Magician |
|  | 79299908 | Singing telegram service |
| Venues | 58130200 | Night clubs |
|  | 58130201 | Cabaret |
|  | 58130202 | Discotheque |
|  | 58129908 | Dinner theater |
|  | 65120300 | Property operation, auditoriums and theaters |
|  | 65120301 | Auditorium and hall operation |
|  | 65120302 | Theater building, ownership and operation |
|  | 79110000 | Dance studios, schools, and halls |
|  | 79110100 | Dance hall services |
|  | 79110101 | Dance hall or ballroom operation |
|  | 79110102 | Discotheque, except those service alcoholic beverages |
| Education | 79110200 | Dance instructor and school services |
|  | 79110201 | Childrens' dancing school |
|  | 79110202 | Dance instructor |
|  | 79110203 | Dance studio and school |
|  | 79110204 | Professional dancing school |
|  | 82990300 | Music and drama schools |
|  | 82990301 | Dramatic school |
|  | 82990302 | Music school |
|  | 82990303 | Musical instrument lessons |
|  | 82990304 | Voice lessons |
| Support services | 59999925 | Theater programs |
|  | 76991000 | Musical instrument repair services |
|  | 76991001 | Organ tuning and repair |
|  | 76991002 | Piano tuning and repair |
|  | 79220000 | Theatrical producers and services |
|  | 79220100 | Theatrical talent and booking agencies |
|  | 79220101 | Agent or manager for entertainers |
|  | 79220102 | Booking agency, theatrical |
|  | 79220103 | Casting bureau, theatrical |
|  | 79220104 | Employment agency: theatrical, radio and television |
|  | 79220105 | Entertainment promotion |
|  | 79220106 | Talent agent, theatrical |
|  | 79220200 | Theatrical production services |
|  | 79220201 | Ballet production |
|  | 79220202 | Community theater production |
|  | 79220203 | Performing arts center production |
|  | 79220401 | Legitimate live theater producers |
|  | 79220500 | Theatrical rental services |
|  | 79220501 | Equipment rental, theatrical |
|  | 79220502 | Scenery rental, theatrical |
|  | 79220600 | Costume and scenery design services |
|  | 79220601 | Costume design, theatrical |
|  | 79220602 | Scenery design, theatrical |
|  | 79229901 | Beauty contest production |
|  | 79229902 | Concert management service |
|  | 79229903 | Lighting, theatrical |
|  | 79229904 | Ticket agency, theatrical |
|  | 89990400 | Songwriting |
|  | 89990401 | Music arranging and composing |
